# Predicting and interpreting COVID-19 transmission rates from the ensemble of government policies

**DOI:** 10.1101/2020.08.27.20179853

**Authors:** C. K. Sruthi, Malay Ranjan Biswal, Brijesh Saraswat, Himanshu Joshi, Meher K. Prakash

## Abstract

Several questions resonate as the governments relax their COVID-19 mitigation policies - is it too early to relax them, were the policies as effective as they could have been. Answering these questions about the past or crafting newer policy decisions in the future requires a quantification of how policy choices affect the spread of the infection. Policy landscape as well as the infection trajectories from different states and countries diverged so fast that comparing and learning from them has not been easy. In this work, we standardize and pool together the ensemble of lockdown and graded re-opening policies adopted by the 50 states of USA in any given week between 9th March and 9th August. Using artificial intelligence (AI) on this pooled data, we build a predictive model (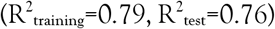, 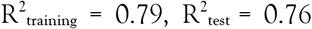) for the weekly-averaged transmission rate of infections. Predictability conceptually raises the possibility of an evidence-based or data-driven mitigation policy-making by evaluating the relative merits of the different policy scenarios. Probing the predictions with interpretable AI highlights how factors such as the closing of bars or the use of masks influence transmission, effects which have been hard to decouple from the ensemble of policy instrument combinations. While acknowledging the limitations of our predictions as well as of the infection testing, we ask the theoretical question if the observed transmission rates in the states were as efficient as they could have been under various levels of restrictions, and if the mitigation policies of the states are ‘overdesigned’. The model can be further refined with a more detailed inclusion of geographies and policy compliances, as well as expanded as newer policies emerge.

## INTRODUCTION

In March-April of 2020, most countries had adopted a partial or complete lockdown in response to the growing COVID-19 pandemic. The lockdowns were mainly motivated by the rapidly rising infections, and a lack of preparedness for the critical healthcare equipment. The added justification came from epidemiological models which projected severe loss of lives for several scenarios of basic reproduction rates (R_0_) characterizing the spread of infections and the case fatality rates [**1,2**]. Subsequent counterfactual models after the lockdowns underscored the reduction in the time varying reproduction rates (R_t_) [**3**] and loss of lives averted by the lockdowns and provided a *post hoc* justification.

Within a month of these partial or full lockdowns, governments began graded re-openings to contain the economic losses. With no vaccines in sight despite intense research efforts [**4**], WHO warned the countries of resurgence if they opened up too soon. As feared, there were resurgences in many countries. Several states and even cities had autonomously implemented restrictive measures over-riding the decisions at the national or state level, and even entering into legal battles on these issues. These autonomous decisions were based on first-hand information about the clusters of infections stemming for example, from night clubs or because of the understanding that masks prevent the dispersion of the droplets.

In the pandemic situation, the immediate attention of the public or the governments concerned about critical healthcare resources is drawn to the number of daily new infections and cumulative infections. However, these infections are a consequence of the transmission or reproduction rates. Several non-pharmaceutical interventions including government policies dictate these rates [**5**]. While this causal relation is implicitly understood and assumed to be true, predicting how the policies translate to the transmission rates has remained a missing link in data-driven or evidence-based decision making (Figure 1). Data driven or evidence-based decision making by exploiting the data and artificial intelligence are being hailed as the next frontiers in many areas, such as public policy making [**6**] or medicine. However, in the COVID-19 pandemic, artificial intelligence had been used mainly for surveillance and for diagnostics, and not for policy making.

**Figure 1.**
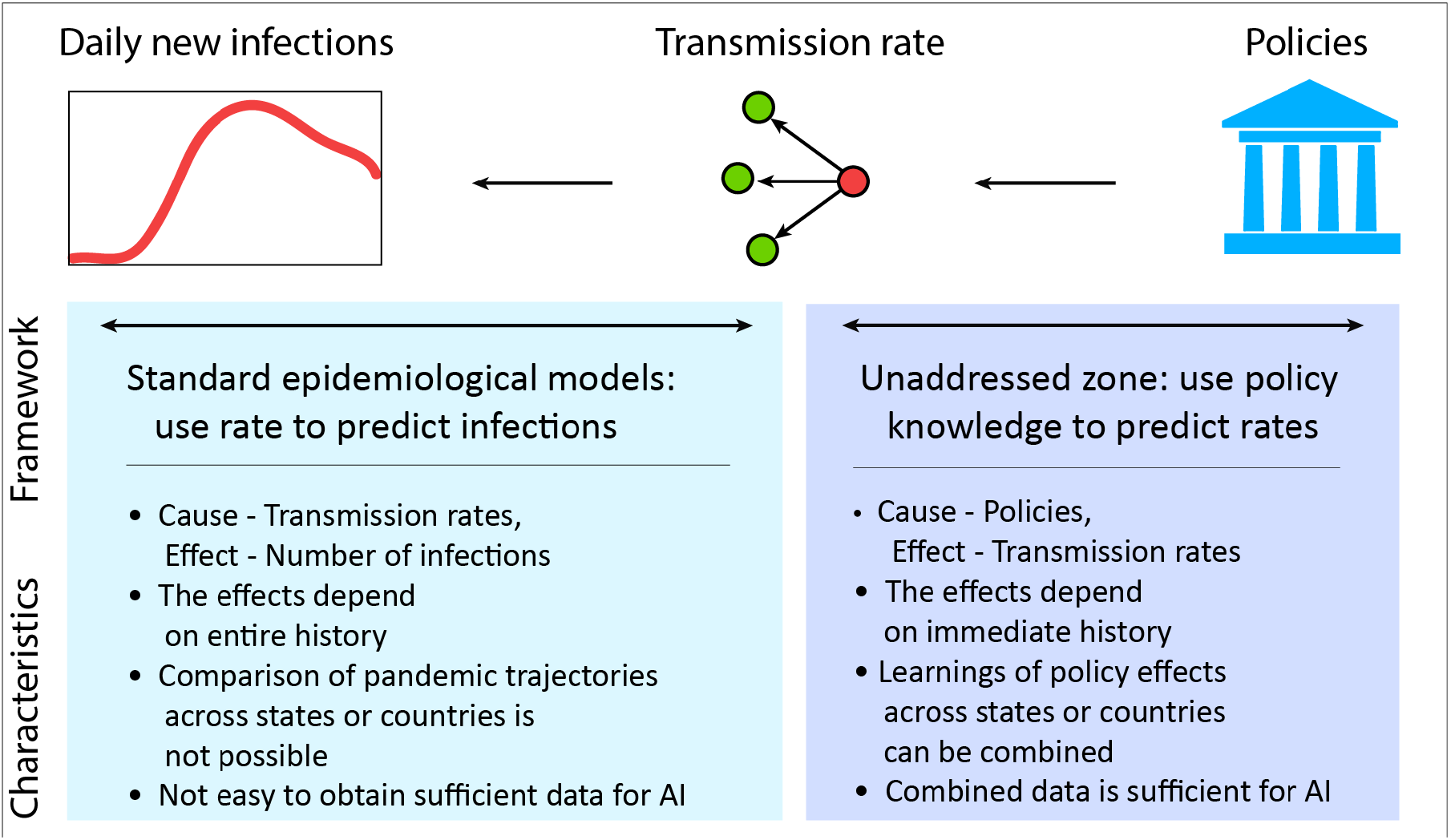
Framework for pooling data on COVID-19 mitigation. While one’s immediate attention is drawn to the numbers of daily cases, they are driven by the transmission rate of infections which in turn could be controlled using non-pharmaceutical interventions such as government policies. Much of quantitative epidemiology focused on the part of predicting infections using rates. However, by focusing on the transmission rates, one can integrate the data from many places into a common framework for learning about the relation between the government policies and the infection rate.

Many epidemiological predictions studied the consequences of different R_t_’s and highlighted the need to keep it in control [**7**]. However, the epidemiology of COVID-19 pandemic had been quite unique [**8**], as it defied the intuitions on infectiousness period, pre-symptomatic and asymptomatic transmission [**9, 10**] of most respiratory infections, including SARS 2003. How the different non-pharmaceutical interventions can reduce the rates to an acceptable level was not clear and each government used a different combination of the different non-pharmaceutical intervention policies. There have been no predictive models translating the policy decisions to the transmission or reproduction rates.

Predicting the rates requires training a mathematical or computational model on sufficiently large and systematized data that contains the information on the policies and their consequences. In the initial days of the pandemic, one could find universalities in the rise of infections across the different countries [**11, 12**]. However, as the policies and the trajectories diverged, this ability to compare or combine data from across the different countries had been lost. A few studies attempted to pool together the learnings by focusing on the reproduction rates which can be compared across different situations. The effectiveness of policy measures from the data pooled from several European countries during February to May [**13**] demonstrated a significant reduction in the reproduction rate due to lockdown. However, many of the interventions in March were implemented in rapid succession and their individual effects could not be decoupled. Other studies reported time-varying reproduction numbers with a detailed state-level tracking in United States [**3**], and correlated them to the mobility patterns [**14**].

In this work, we build a predictive model by pooling together the policy and infection data from the 50 states of United States in 9 March - 9 August period. Through this period, these states witnessed prelockdown and lockdown phases along with a diverse combinations of policy instruments ranging from a graded re-openings of restaurants, bars, to imposition of mask mandates. An implicit assumption is that the different states of the USA are similar in many ways including testing policies and rigor of policy implementation, and that there is an opportunity to learn from other states which experienced same or similar situations. The model which is developed using the data from different states can of course be refined on a continuous basis by also including the standardized data from different countries.

The aim of this work is to introduce a data-driven framework for COVID-19 mitigation with three elements: firstly, to show that the transmission rates can be predicted from the knowledge of standardized policies, secondly demonstrate the possibility to decouple the relative contributions of the individual policy instruments, and finally to understand if the policies are overdesigned by checking hypothetical relaxation of policies in the model.

## RESULTS AND DISCUSSION

### Standardization for data-driven policies

A data-driven approach will require a standardized set of the different non-pharmaceutical instruments and their measurable consequences. For the latter, we use the transmission rates, which depends on how people interact with each other and is independent of number of current number of infections. Most state governments of USA used various combinations of the policy instruments such as the degree of opening of the retail stores, restaurants, bars, personal care businesses such as barber shops, nightclubs, child care, places of entertainment such as movies, allowed size of gatherings, places of worship, beaches along the oceans or lakes, and outdoor activities. The policy of the states on quarantine restrictions upon inter-state arrivals, and the rigor of mask requirement were also noted and used as independent variables. 14 different policy instruments and the average mobility from Google [**14**] each week were used as the independent variables. These degrees of opening were codified as discussed in the **METHODS** section. For example, the codes we assigned to the restaurant opening policies were: 0 (completely closed), 1 (allowed for a takeaway), 2 (allowed up to 25% seating capacity), 3 (allowed up to 50% of seating capacity), 4 (allowed for more than 50% seating capacity), 5 (pre-pandemic occupancies). A true pre-pandemic situation, with a 100% capacity and no frequent hand washes or use of personal sanitizers, may not be reached in the near future.

The government policies on graded closing or re-openings were gathered from the press releases of the Governors of the 50 states of USA. We discretized the period under study into weeks. For simplicity we follow the standard week numbering convention that identifies the week starting 9^th^ March 2020 as the week 11. If a policy change was implemented anytime during the week, the code for the new policy was uniformly applied throughout the week.

In reality, a restaurant may not be populated, despite a relaxation of the restrictions and additional data will be required to understand these subtle differences between allowance and compliance. However, in this work we assigned the codes to all the independent policy variables based on the limits allowed by the governments. Whether or not the restrictions of gatherings were applicable to the places of worship was not considered. Also, during this period starting from spring break till the summer, schools were closed throughout the United States and hence it was not considered either in our present model, although newer situations and policies may be added in future.

### Transmission Rates

Reproduction ratios are standard epidemiological descriptors, which clarify the relative rates of transmission (α_t_) and recovery from the infection (γ). However, the emphasis of the present work is on transmission alone. We assume simple first order kinetics that the new cases depend on the currently active infections and the rates. The daily transmission rates were calculated as 1/I_active_. (dI/dt) where I_active_ is the number of active infections, and dI/dt is the number of daily new infections on a given day. The daily new infection data was obtained from Worldometer [**15**]. We work with the assumption that the function is piece-wise continuous [**13**], with the resolution of a week [**12**]. The average transmission rate for the week was calculated, α_t_=<1/I_active_. (dI/dt)>.

The instantaneous daily rates, their weekly averages α_t_ and how they change over the weeks is illustrated in Figure 2 and demonstrates a dynamic scenario. Assuming constant recovery rates, the transmission rates we calculate can be correlated to the reproduction rates [**3**]. The results in this work are mainly reported as the transmission rates (α_t_). However, in some parts of this work, time dependent reproduction ratios were estimated from the transmission rates (R_t_=20 α_t_), by using the average recovery time of 20 days (γ=1/20) between an overall 24 days [**16**] and the 15 days for people requiring medium-care [**17**] and that have been reported.

**Figure 2.**
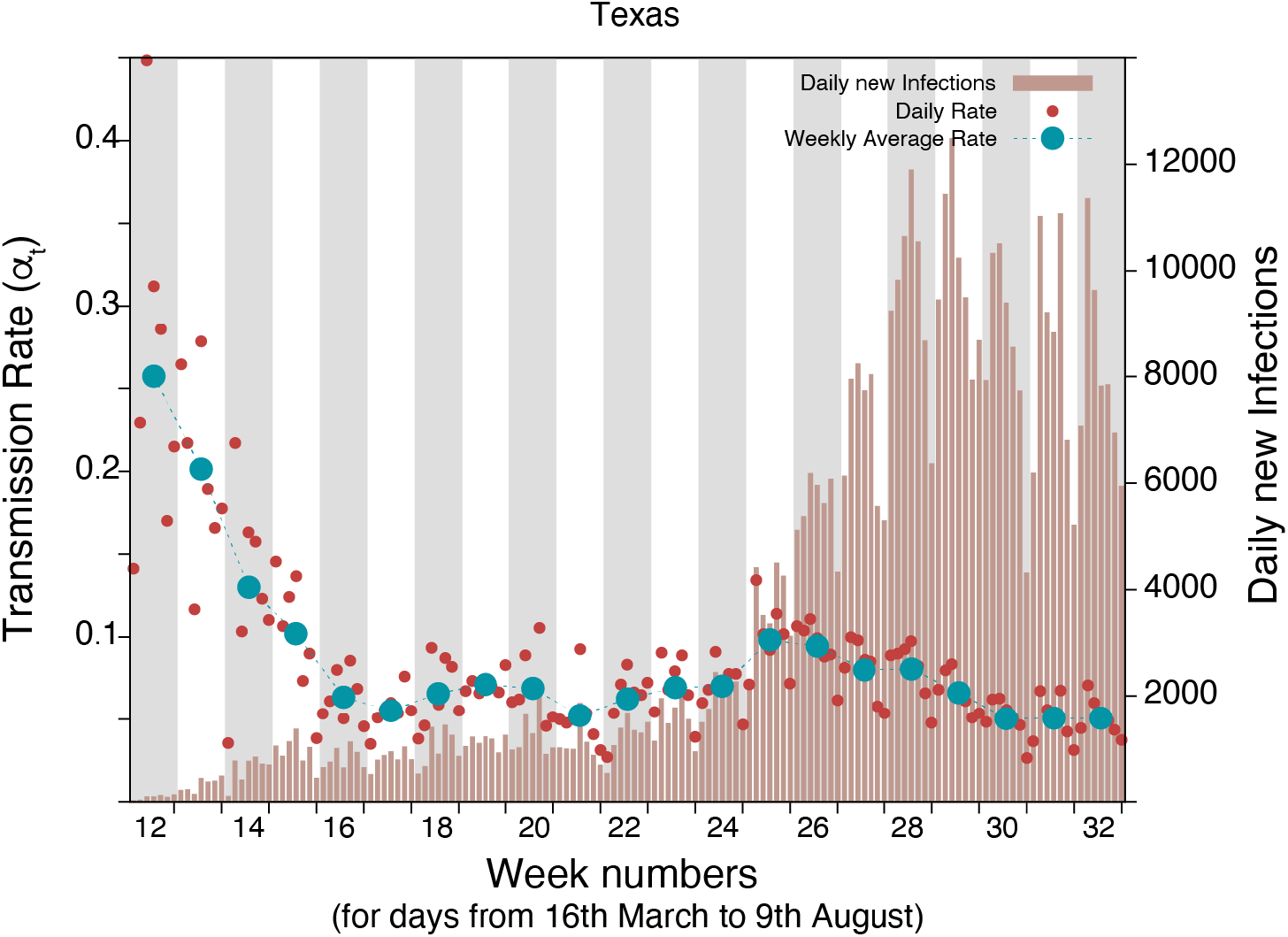
Daily infections and transmission rates. The graphic shows the daily new infections reported by Texas between 16^th^ March (week number 12) and 9^th^ of August (week number 32). Also shown are the daily instantaneous rate 1/I_activeis_·dI/dt as well as its weekly average α_t_= <1/Iactive·dI/dt > that we calculated. The rate that is observed in a week is assumed to be a result of the policies in the earlier week.

### Machine learning Model to Predict Transmission Rates

Our goal in this work is to show that the transmission rates or alternatively the reproduction rates can be predicted from the policy decisions, by learning from same or similar experiences from all states. Considering a median incubation period of around 5-6 days [**18**], the policies adopted in a week are assumed to affect the transmission rates observed in the following week. The weekly policy data from 50 states from 9 March (Week 11) to August 2 (Week 31) were used as independent variables to predict the observed rates in the weeks 12 to 32. We developed an artificial intelligence model based on XGBboost for this purpose (**METHODS**). 14 different policy instruments and the average mobility from Google [**14**] each week were used as the independent variables. The hyperparameters were optimized using the RandomSearch, and calculations performed in Python (**METHODS**).

Seven states never issued a stay-at-home order, although businesses were closed. Others did so at different times between 16 March-31 March, while beginning some recommendations or restrictive measures. Thus, it was not easy to codify the policy data especially until March 31. We thus performed the calculations with two scenarios, one assuming the states were not applying restrictions until they announced the stay (or safe) at home order, and another from the day the graded shutdown recommendations were made in mid-March. Using either of the data sets we obtained similar results. In both cases we trained the model on 80% of the 1071 data points from 51 states and 21 weeks and tested it on the remaining 20%. We obtained comparable results (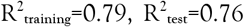 for both), indicating that predictions of the rates are robust to some differences in interpretation of policy. The complete set of results are in the **Supplementary Information**. In this work we only discuss the first scenario. The values of observed and calculated rates (Figure 3A) or their patterns over the weeks correlate well (Figure 3B). The model we developed using the policies adopted by the different USA states demonstrates the predictability of the transmission rates from the knowledge of the policies. Further, we believe the approach which predicts the transmission rates to be observed in the following week also implicitly suggests how quickly one can expect to see the results of major policy changes.

**Figure 3.**
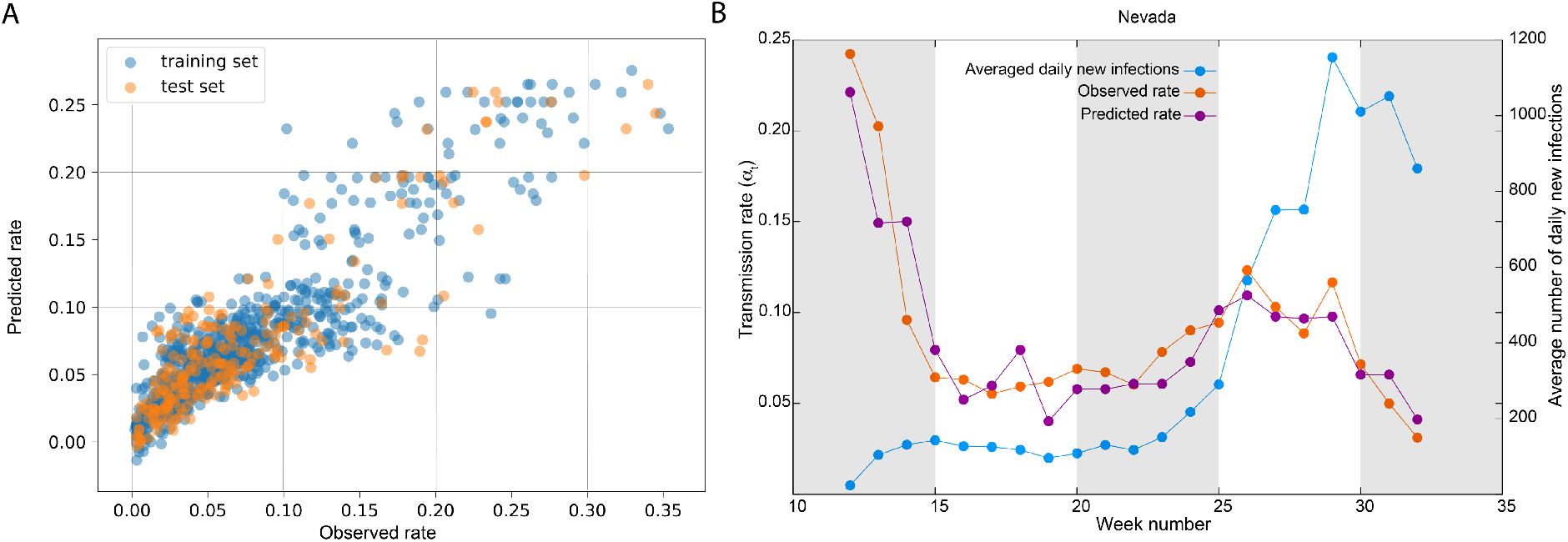
Correlations between predicted and observed rates. A. The overall quality of the training and test data (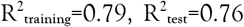) is shown. B. The weekly pattern of observed rates and predicted rates. The average number of daily new cases in that week is also shown for clarity.

### Decoupling the Contributions from the Different Policy Instruments

We probed the model to understand the contributions of the individual policy instruments such as the closure of restaurants or enforcement of masks to the prediction of rate of every week, from every state. Since various combinations of these different policy instruments were used it may not be easy to decouple them. We used the interpretable AI framework based on SHapley Adaptive exPlanation (SHAP) for this purpose [**19**]. SHAP values are the linear contributions from each variable to every prediction, a negative number signifying a contribution to the reduction in the transmission rate. A summary comparing these SHAP contributions from the different variables (Figure 4), shows the range of contributions that arise from each policy instrument or feature. Some of the highest contributions to the overall set of predictions come from the graded openings of bars, restaurants as well as the imposition of mask mandates (Figure 5A, 5B, 5C). Mobility data [**14**] was used by several governments during the stay-at-home period to track the compliance to the orders. The average movement of people relative to the baseline correlated with the reproduction rates during the lockdowns [**3**]. However, in our calculations, where various degrees of re-opening are also included, average mobility does not show any correlation with the rates (Figure 5D).

**Figure 4.**
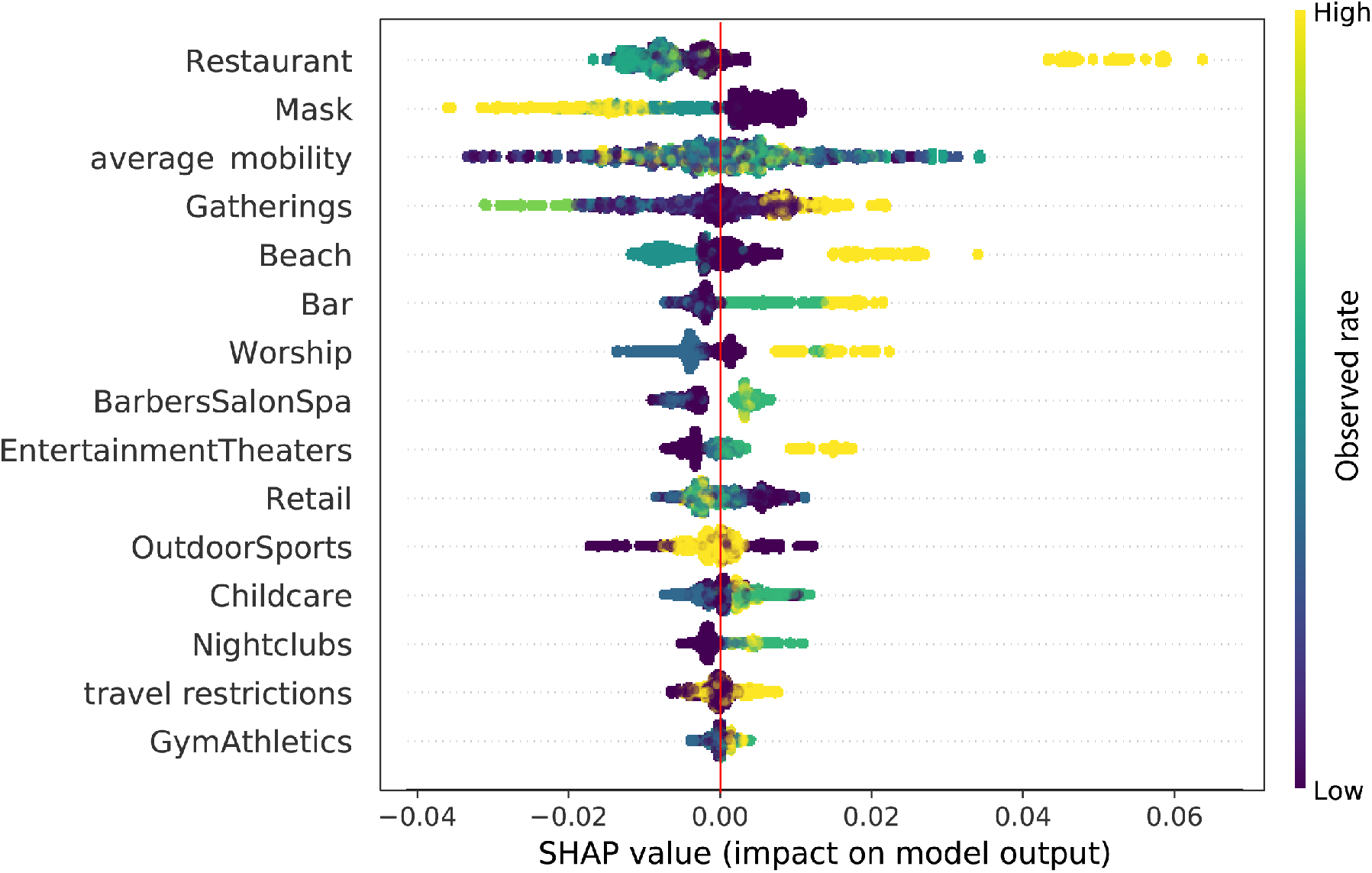
Contribution from individual factors. We used the SHAP interpretable AI framework to identify the contributions from individual variables to the transmission rates, at. The color bar indicates the relative value of the individual policy instrument or feature. Restaurants, and masks emerge as dominant features. Mask is anticorrelated (as can also be seen in Figure 5C) because the enforcement of masks (code 2 in our scheme) reduces the spread compared to no-mask scenario. Mobility has high contributions in magnitude, but no correlations could be established. Separating these contributions is an attempt to decouple the different factors using the tools from AI, but the relation cannot be immediately interpreted as being causal.

**Figure 5.**
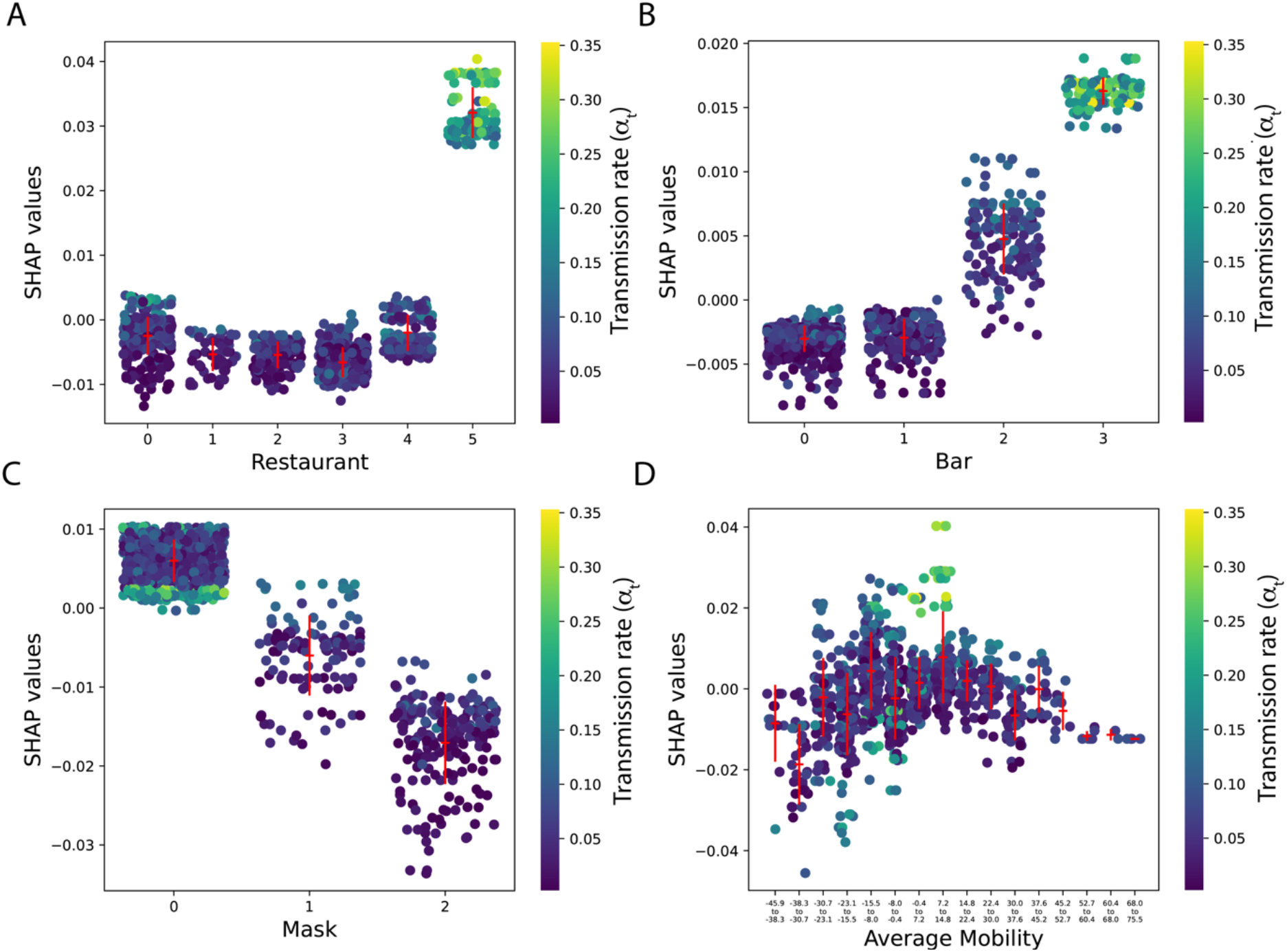
Correlations of factors to transmission rates. The SHAP contributions of the individual variables to the individual rate predictions are shown for four different variables. The mean and standard deviation of these categories are shown in red. A significant difference beyond the errorbar is seen in only some cases. In our analysis, Mobility information did not display any significant differences, despite high contributions. Here the colorbar indicates the transmission rate α_t_.

Another advantage of SHAP is that it separates the contributions of various factor to each single prediction. For example, Figure 6 illustrates the different factors contributing to the transmission rates in Alabama in week 28, 29. One can clearly see that the role of the Mask variable changed from increasing the rates (week 28) to reducing the rates (week 29), the week in which mask mandate was introduced in Alabama. The causal relations cannot be over interpreted. However, the purpose of the work was to demonstrate the possibility of such interpretations with larger data, and possibly with tools better suited for the same.

**Figure 6.**
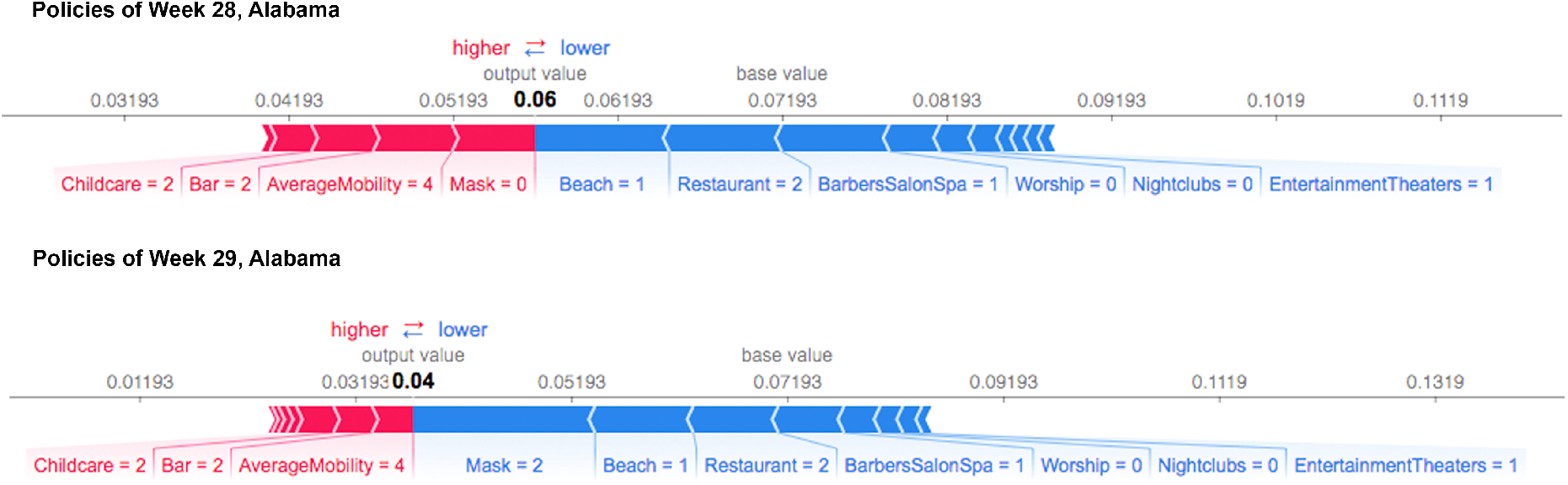
SHAP decomposition of individual contributions. The transmission rate in Alabama changed from 0.06 in week 28 to 0.03 in week 29 (R_t_ from 1.2 to 0.6 assuming γ=1/20). Mask mandate was introduced on July 16 (Week 29). We used SHAP framework to decompose the contributions of individual factors to the transmission rate prediction. It can be clearly see that the role of the Mask switched

### Benchmarking the observations and understanding overdesign

Ability to predict the efficacy of policy measures may also serve two additional purposes: to benchmark and understand if the policy was as effective as it could have been and also to understand if the policies have been overdesigned. We explore these conceptual questions, by *assuming* the model to be reliable in estimating the transmission rates. The difference between the observed and predicted rates was categorized into three groups using R_t_ (R_t_ < 1, 1 < R_t_ < 2 and 2 < R_t_). Our analysis shows that when the rates are low, the observed rates and the expected one based on a model trained on all states correlate well. However, at higher R_t_ values, there are some differences. While the differences clearly indicate that the model has to be improved, the possibility that the reported infection data may not be accurate due to inefficient testing or delayed reporting cannot be dismissed.

Drawing upon the notions from engineering, we consider a policy to be overdesigned if relaxing some of the constraints does not affect the desired outcome significantly. This analysis was performed by relaxing the constraints on each of the individual policy instruments to their next natural step and noting the difference in the transmission rates. For example, restaurant code 2 was relaxed to 3, indicating the change from a takeaway policy to a 25% occupancy at the restaurant, and the change in rate relative to the baseline value of that week as predicted by the model is noted. Figure 7 shows the overall distribution of the changes of rates when one policy is relaxed or when any two policies are relaxed. According to these results for week 31 predicted by our model, the policies of California are overdesigned and those of Missouri are most sensitive to relaxation. Whether this relaxation of one or two instruments is acceptable depends upon the tolerance of the state to the number of new cases that may arise, but such knowledge may help in planning the relaxation of containment policies.

**Figure 7.**
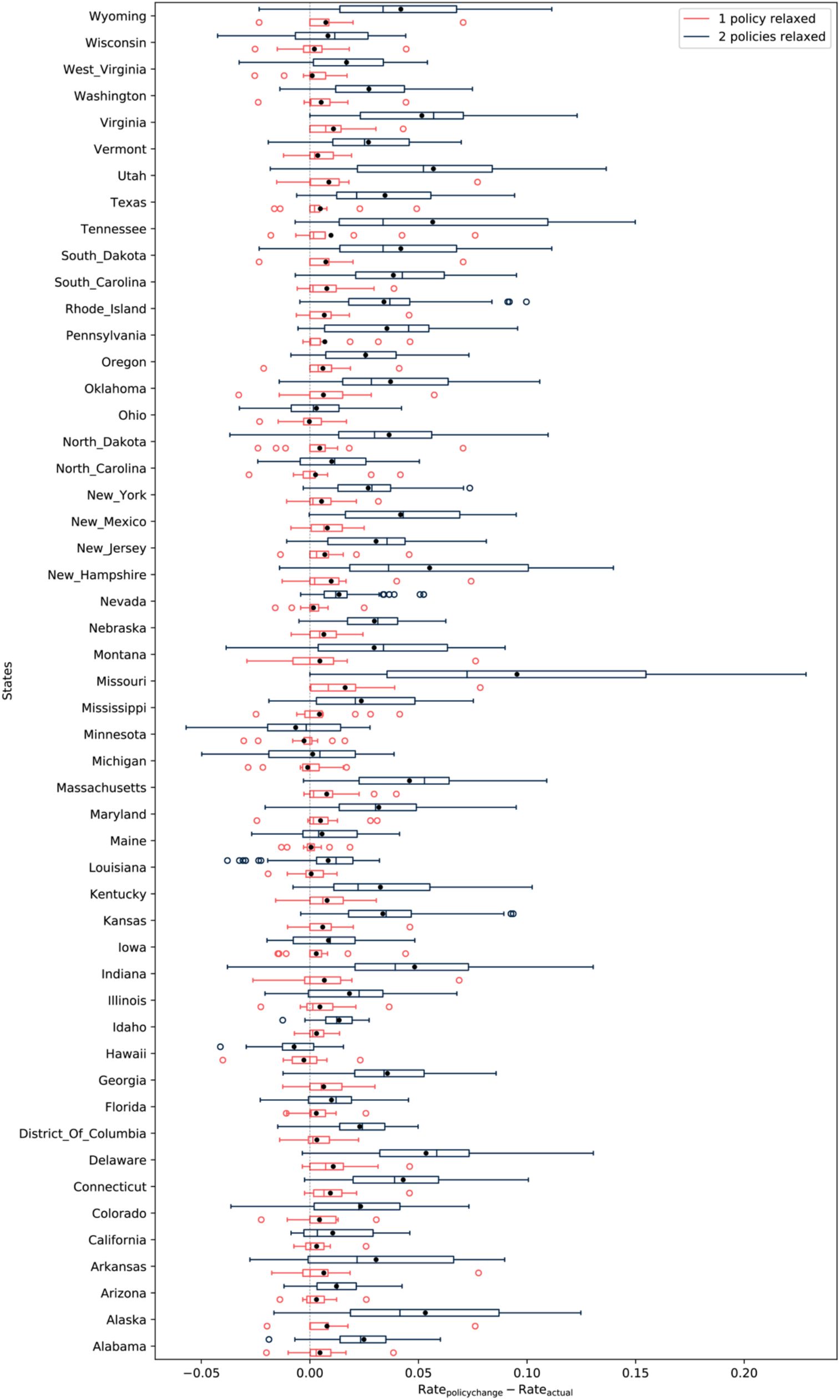
Overdesign. If the conditions are overdesigned for the baseline transmission rate that is currently active, then relaxing one or two constraints will have minimal effect on the rate. The rate data for week 31 is shown. The effect of relaxing one or any two constraints on the baseline transmission rate is examined It is clear that relaxing two constraints increases the transmission rate.

### Integrating More Data from Newer Scenarios

The conceptual illustration of the overdesign of policies above uses our model predictions as a reference. For the model to serve as a gold standard, it needs to be trained on more data from newer geographies and newer scenarios. The ensemble of policy measures which were unguided initially, can now begin to be structured using the evidence that is building up from many countries and states. The policies adopted by the states of USA had been quite different. Although most of them responded to the national emergency with lockdowns, the reopening strategies were quite different. However, purely from the modelling perspective the ensemble of policies the governments adopted help in training better models. Fine-graining the data from different counties in a state, and following their individual policies and rates will serve the dual purpose of removing regional heterogeneities in rate estimation and increasing the data available for the analysis. Integrating data from other countries which may have experienced newer scenarios like the opening of the schools will be helpful and can be done after standardizing it for the rigor of implementation of the policies, and testing. Needless to say, a similar understanding of the economic losses associated with the policies are also required for fully informed decisions.

### Conclusions

In summary, we show that a predictive model could be built by integrating the data from different locations, despite the differences in the policy landscape. Standardizing the data from more geographies, using refined tools of AI, one will be able to improve the predictions and interpretations about the role of the policy instruments on the COVID-19 transmission rates, providing guidelines for a data-driven policy-making.

## METHODS

### Policy data curation

The information about the policy data was curated from the official press releases of the Governors of the states and by cross-checking them with the media reports. The following codes were employed:

- *BarbersSalonSpa:* Everything closed (0), Beauty salons, Barbers opened (1), nail spas, tanning salons, tattoo parlors also allowed to operate (2), Prepandemic level (3)
- *Bars:* Bars are places which primarily sell alcohol and not food. Closed (0), Opened with 25% capacity (1), Opened with 50% capacity (2), Pre-pandemic level (3)
- *Beaches*: Beaches on oceans such as in California or Lakes in states without ocean but still attracting tourists were considered. Beaches closed (0), open with severe restrictions (1), open with lesser restrictions (2), Pre-pandemic level (3)
- *Childcare:* Closed (0), Open (1), Summer camps allowed as well (2), Pre-pandemic level (3)
- *Gathering size:* The allowed gathering sizes were used - 0, 3, 5, 9, 10, 25, 50, 100, 200, 250 (when it was mentioned as 250 or unrestricted)
- *Mask*: No mask requirement (0), Mask usage is recommended but not mandatory or mandatory in some cities (1), Mask usage is mandatory (2)
- *Mobility*: Mobility data relative obtained from Google COVID-19 Community Mobility [**Google**] Reports, and was used as an average of all the categories that were given
- *Nightclubs:* Closed (0), Opened with 25% capacity (1), Opened with 50% capacity (2), Pre-pandemic level (3)
- *Outdoor sports:* Stay at home with no outdoor activities (0), Parks, tennis courts, dog parks, hikes allowed and also Pre-pandemic level (1)
- *Restaurants:* Closed (0), Takeaway (1), fractional capacity of up to 25% (2), fractional capacity of up to 50% (3), Unrestricted (4), Pre-pandemic level (5)
- *Retail*: All non-essentials closed (0), partial opening (1), extended opening including books, florists, etc. or auto showrooms (2), Pre-pandemic level (3)
- *Travel restrictions*: No quarantine restrictions upon interstate arrival and also Pre-pandemic level (0), Inter-state arrivals from some or all states are asked to quarantine (1)
- *Worship*: Places of worship such as Churches closed (0), open with restrictions (1), Pre-pandemic level (2)

There were other state specific parameters such as opening of movie shootings in California, high spread of infections in the meat industry in Nebraska. All these state specific parameters were not considered. The number of tests conducted could be another variable. However, we did not use it in the present model.

### AI model

We used the *XGBoost* library in Python for developing an AI model to predict the rate. 80% of the complete data set consisting of 1071 data points, was used as the training set and 20% as the test set. The parameters *learning_rate, n_estimators, max_depth, min_child_weight, gamma, colsample_bytree, subsample* and *reg_alpha* were tuned using *RandomizedSearchCV* function. *RandomizedSearchCV* creates n-sets of parameters (n=1000 in our case) with each parameter value being selected from the list of different values that are to be considered for tuning. Models are developed using these parameters by training on the chosen data set. Each model’s predictive ability is evaluated using a 5-fold cross-validation analysis and the best set of parameters is selected based on the average of mean squared error for the validation sets. The parameters that yielded the lowest validation error for our data set were *learning_rate=0.04, n_estimators=1300, max_depth=3, min_child_weight=15, gamma=0, subsample=0.8, colsample_bytree=0.85, reg_alpha=0.0001*. With these parameters, the RMSEs for the training and test sets were 0.0279 and 0.0320, respectively. The quality of prediction seemed to be robust against changes in the training set as shown by the 5-fold cross-validation analysis, with a standard deviation of 0.0005 and 0.0029 for the training and the test set RMSEs.

### Individual variable contributions

Contribution of each variable towards each prediction was obtained through SHAP analysis of the model with the tuned parameters. The Python implementation of SHAP (https://github.com/slundberg/shap) was used for our interpretable AI analyses as well as for generating figures. The analyses were repeated with a 5-Fold cross-validation and the differences were found to be minimal.

## Data Availability

The work is a theoretical work performed with publicly available data on the number of infections. However, the data will be made available upon request.

## Acknowledgements

We thank Dr Hari Thadakamalla for helpful discussions.

## Author contributions

CKS, MRB performed the machine learning calculations; MRB, HJ, BS extracted the epidemiological data and performed analyses of rates; CKS, MRB, HJ, BS, MKP analyzed the results; MKP conceived the project, curated the data on government policies and wrote the paper;

## Supplementary Data Availability

The source data, and figures for all states are available at https://github.com/meherpr/COVIDrates

## Competing interests statement

The authors declare no competing interests.

---

**Supplementary Figure 1.**
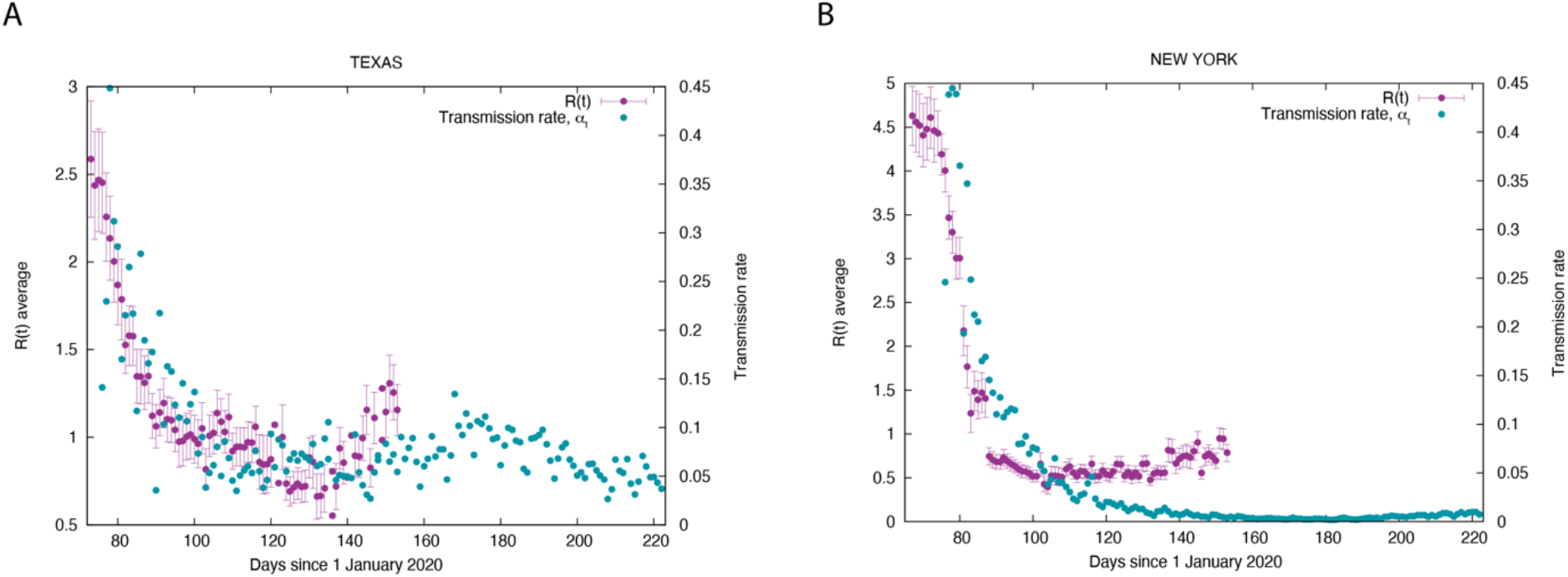
A comparison of the transmission rates (α_t_) calculated in this work with the reproduction rates (50% CI) obtained from Unwin et al. [**3**].

## Notes

### Competing Interest Statement

The authors have declared no competing interest.

### Funding Statement

No funding available

### Author Declarations

The work is a theoretical work performed with publicly available data on the number of infections. No ethics approval is required.

